# Wildlife hosts predict the distribution of reported coccidioidomycosis in the western United States

**DOI:** 10.64898/2026.03.10.26348058

**Authors:** Jacob Sussman, Katrina M. Derieg, Kevin D. Perry, Ashlyn Adakai, Raniah Corrian, Cory Merow, Simon C. Brewer, Katharine S. Walter

## Abstract

Global environmental change is reshaping human exposure to zoonotic and environmentally acquired pathogens, yet predicting disease risk remains challenging. High-resolution risk maps typically rely on human case data and environmental correlates, often overlooking ecological processes such as wildlife reservoirs. We evaluated whether mammalian reservoir distributions improve prediction of coccidioidomycosis (Valley fever), an emerging, environmentally-acquired fungal disease with a poorly characterized range. Using county-level coccidioidomycosis notification data from the Centers for Disease Control and Prevention, we developed a hierarchical Bayesian model of county-level endemicity, defined as >=10 cases per 100,000 population. We incorporated climatic, environmental, and vegetation covariates, a state-level reporting effect, and species distribution models for 22 mammalian species previously identified as *Coccidioides* reservoirs. We found that the number of endemic mammalian reservoirs in a county was the strongest predictor of coccidioidomycosis endemicity, with each standard deviation increase in reservoir species richness associated with substantially higher odds of endemicity (log-odds ratio = 1.702; 95% CI: 1.060-2.419). In contrast, maximum vapor pressure deficit, soil moisture, and land cover were not independently associated with endemicity after accounting for reservoir distributions. State-level reporting effects revealed substantial heterogeneity, and comparison of models with and without reporting effects identified regions likely to be endemic but underreported, including parts of Nevada, Utah, New Mexico, Texas, and Colorado. Our results establish reservoir diversity as a central predictor of zoonotic fungal disease risk and demonstrate a transferable framework for distinguishing between ecological drivers of infection from surveillance bias to improve disease risk mapping and identify areas of potential underreporting.

## Introduction

Zoonotic and environmentally acquired pathogens are responsible for a large proportion of human infections and are among those most sensitive to global change^1,2^. Unlike for many directly transmitted human diseases, vaccines are often unavailable and public health measures must focus on preventing zoonotic and environmental exposures. Yet we have only a limited understanding of the environmental distributions of many pathogens due to the prohibitive costs of systematic surveys, and must rely on reported human case data, which is sparse and biased.

Coccidioidomycosis, or Valley fever, a fungal disease caused by inhalation of *Coccidioides immitis* and *C. posadasii* spores, is one such disease with poorly defined infection risk. Systematic environmental sampling for *Coccidioides* is infeasible. Therefore, skin testing studies conducted in the 1950s^3,4^, along with contemporary reported cases of disease remain the basis for the Center of Disease Control and Prevention’s current Valley fever risk maps^5^. Improved risk maps are urgently needed because reported cases of the disease in the United States have dramatically increased in recent years, from 2,271 in 1998 to 21,037 in 2023^8^. Poverty, outdoor occupations such as farming and construction, and incarceration in endemic areas exacerbate risk, compounding inequities in healthcare access ^6^.

Although climatic and soil characteristics correlate with fungal range, they are insufficient predictors of risk of disease^7^. Further, while soil has long been considered the primary reservoir of *Coccidioides spp.*, there is increasing evidence for the endozoan hypothesis, that zoonotic hosts maintain *Coccidioides spp.* and seed the soil with infectious spores when they die^8^. Biodiversity data is increasingly available^9,10^, and host distributions have been successfully incorporated into risk mapping for several zoonotic diseases, including Lassa fever, plague, and hantaviruses, where the presence or abundance of small mammal reservoirs strongly predicts human spillover. Building on this precedent, we tested whether small mammal distributions help explain the endemicity of *Coccidioides*, thereby providing a novel ecological layer for mapping Valley fever risk.

Coccidioidomycosis case reports are subject to substantial biases, making it difficult to infer the true distribution of endemicity using standard modeling approaches. Many infections are never reported because symptoms are mild or resemble other respiratory diseases, and because of low levels of population and physician awareness and lack of access to healthcare. The true incidence has been estimated to be up to thirty times higher than reported cases^11,12^. Reporting also varies widely across the western United States. Although coccidioidomycosis is a nationally notifiable disease with a standardized case definition, states are not obligated to submit reports to the Centers for Disease Control and Prevention. As a result, several endemic states, including Texas, do not report cases^13^. Beyond reporting biases, infection risk for environmentally acquired pathogens such as *Coccidioides* is spatially autocorrelated, further complicating inference with conventional regression approaches.

Integrated Nested Laplace Approximation (INLA) approaches enable efficient Bayesian spatial modelling of spatially autocorrelated data to generate maps of infection risk^10,14^. INLA models have been successfully applied to map other environmentally acquired and zoonotic pathogens, including leptospirosis^15^, schistosomiasis^16^, dengue^17^, and Zika virus^17^. This approach is therefore well suited to generate updated, data-driven maps of coccidioidomycosis endemicity while rigorously testing ecological hypotheses such as the role of rodent reservoirs.

Here, we developed an INLA model to predict coccidioidomycosis endemicity in the western U.S. We tested whether mammalian reservoir distributions predict coccidioidomycosis endemicity, while also assessing environmental and reporting factors. Our approach refines coccidioidomycosis risk maps and provides a generalizable framework for mapping environmentally acquired pathogens.

## Methods

### Human case data

We obtained Valley fever human case data reported to the National Notifiable Diseases Surveillance System through a data use agreement with the Centers for Disease Control and Prevention (CDC). Cases are reported from 1998 to 2022 by states. In this study, we use the data reported for the year 2022. The data reports a monthly case frequency grouped by state and county Federal Information Processing Standards code. Additionally, the reporting month is identified as the month of disease onset or diagnosis. We summed the total cases reported in each county for each month to obtain the total number of Valley fever cases reported in each U.S. county in 2022. While coccidioidomycosis is a Nationally Notifiable Disease, reporting varies by state, we used the CDC list of reportable diseases to identify states that report coccidioidomycosis^18^.

To align with prediction goals specified by the CDC, we define counties as endemic if incidence is ≥10 cases per 100,000 people, and treated endemicity as a binary outcome. While we are primarily interested in results for counties west of the Mississippi (the “western” U.S.), to avoid introducing artificial spatial boundaries, we include all U.S. counties in our analysis. Regions outside of the spatial neighborhood domain like Alaska, Hawaii, and other territories were not included in analysis.

### County data

We obtained county shapefiles from U.S. census data and accessed through the R package tidycensus^19^. Using the same package, the 2022 county population size was obtained from the American Community Survey^20^.

### Bioclimatic covariates

We obtained bioclimatic data for covariates that have been associated with or that we predicted were associated with *Coccidioides* presence^7,21–24^. We obtained monthly values of maximum temperature, mean temperature, minimum temperature, precipitation, minimum vapor pressure deficit (VPD) and maximum VPD from the PRISM datasets and extracted mean county raster values averaging over all months in 2022 using the R package prism^25,26^. Climate factors like temperature and precipitation are commonly used in models of *Coccidioides* endemicity due to their influence on fungal growth.^7^

We obtained modeled soil properties at a 30-second spatial resolution, and a depth of 0-5 cm from SoilGRIDS, accessed through the package geodata^27,28^. We accessed soil temperature and soil moisture from the 2022 NLDAS-NOAH products from NASA^29^. Soil sampling studies suggest that *Coccidioides* relies on soil as an exclusive source of nutrients, and therefore soil parameters should be considered when building distribution models.^23^

We accessed land cover data from the Multi-Resolution Land Characteristics Consortium database provided by the U.S. Geological Survey.^29^ We assigned each county a single value, the “dominant cover type,” by identifying the most common land cover category raster value in each county. When the dominant land cover type was water, we assigned the next most common cover type. We extracted mean county values for all covariates and attached all data to a simple features (sf) object in R^31^. While the role of land cover and agriculture on *Coccidioides* presence remains unclear, we include land cover covariates in our modeling to strengthen the diversity of ecological factors used outside of mammal distributions.

### Mammal distributions

To test whether reservoir distributions were associated with disease endemicity, we created a map that included the distributions for any mammal previously reported to be infected with *Coccidioides*.

First, we compiled all previous studies documenting *Coccidioides* infection through culture, post-mortem serology, or ITS2 amplicon sequencing of mammal lung tissue ^32–34^ (Supplementary Table 1). We then generated species distribution models (SDMs) for each species, following previously published methods^35^. For each SDM, we converted continuous model outputs into a binary outcome by considering any raster values greater than or equal to 6 to mean that the mammal species is present. We summed the number of species with ranges intersecting each county to obtain a total number of reservoir hosts occurring in each county.

### Covariate selection

We excluded covariates that were highly correlated (>80% correlation). Candidate models were compared using the Widely Applicable Information Criterion (WAIC) and the Deviance Information Criterion (DIC). Candidate models with different covariate subsets were compared using WAIC to identify the final specification. In our final model, we included population, mean vegetation, mean soil moisture, mean soil bulk density (BDOD), maximum VPD, and a simplified grouping of land cover classes, and number of potential reservoirs.

### INLA modeling

We modeled county level endemicity using a Bayesian spatial model fitted with integrated nested Laplace Approximation (INLA v 25.6.7) for areal data. The outcome, a binary indicator of endemicity, was modeled with a binomial likelihood and logit link. To capture county-level spatial autocorrelation we used the BYM2 parametrization of the Besag-York-Mollie model^36^, which describes unexplained county-level variation as the mixture of a spatially-structured intrinsic conditional autoregressive component (𝑢) and an unstructured independent and identically distributed (i.i.d.) component (𝑣):

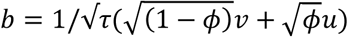

The spatial structure was defined using a county-adjacency matrix based on Queen’s case contiguity. We specified a penalized complexity (PC) prior for the spatial hyperparameters^37^. The prior on the precision (𝜏) was given as 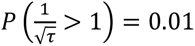, and the prior on the mixing parameter (𝜙) was given as 𝑃(𝜙 > 0.5) = 2/3 ^38^. The PC prior penalizes deviation from a base model with no spatial dependence.

In addition to county spatial effects, we included an i.i.d. state-level random effect to account for unobserved state-specific heterogeneity not captured by the county structure. To characterize the underlying spatial signal in the outcome, we first fit a spatial-only baseline model that included no covariates. The baseline model allows us to evaluate how much of the variation in endemicity is attributable purely to spatial dependence.

We then fit a full covariate model with both county and state-level spatial effects. Fixed effects included population, species presence count, mean vegetation, mean soil moisture, mean soil bulk density, VPD maximum and a simplified land-cover category. We standardized continuous predictors prior to modeling. Models were fit with INLA, and we describe model fit using deviance information criterion (DIC) and Watanabe-Akaike information criterion (WAIC). We obtained the county-level probability of endemicity from the full INLA model by extracting the mean of the posterior fitted values for each county and assigning this value as the probability of endemicity.

To isolate the environmental component of the model, after fitting we extracted the fixed-effect estimates and constructed a design matrix containing only the environmental covariates. We computed the linear predictor corresponding to these fixed effects. The environmental-only prediction reflects the contribution of environmental factors to endemicity independent of county or state spatial effects.

All code is publicly available on GitHub: https://github.com/sussmanjacob/vf_distribution_modeling.

### Human subjects

The University of Utah Institutional Review Board determined this study was Non-Human Subjects Research (IRB_00177442).

## Results

In 2022, 17,562 cases of coccidioidomycosis were reported in the United States (Fig. 1a).

**Figure 1:**
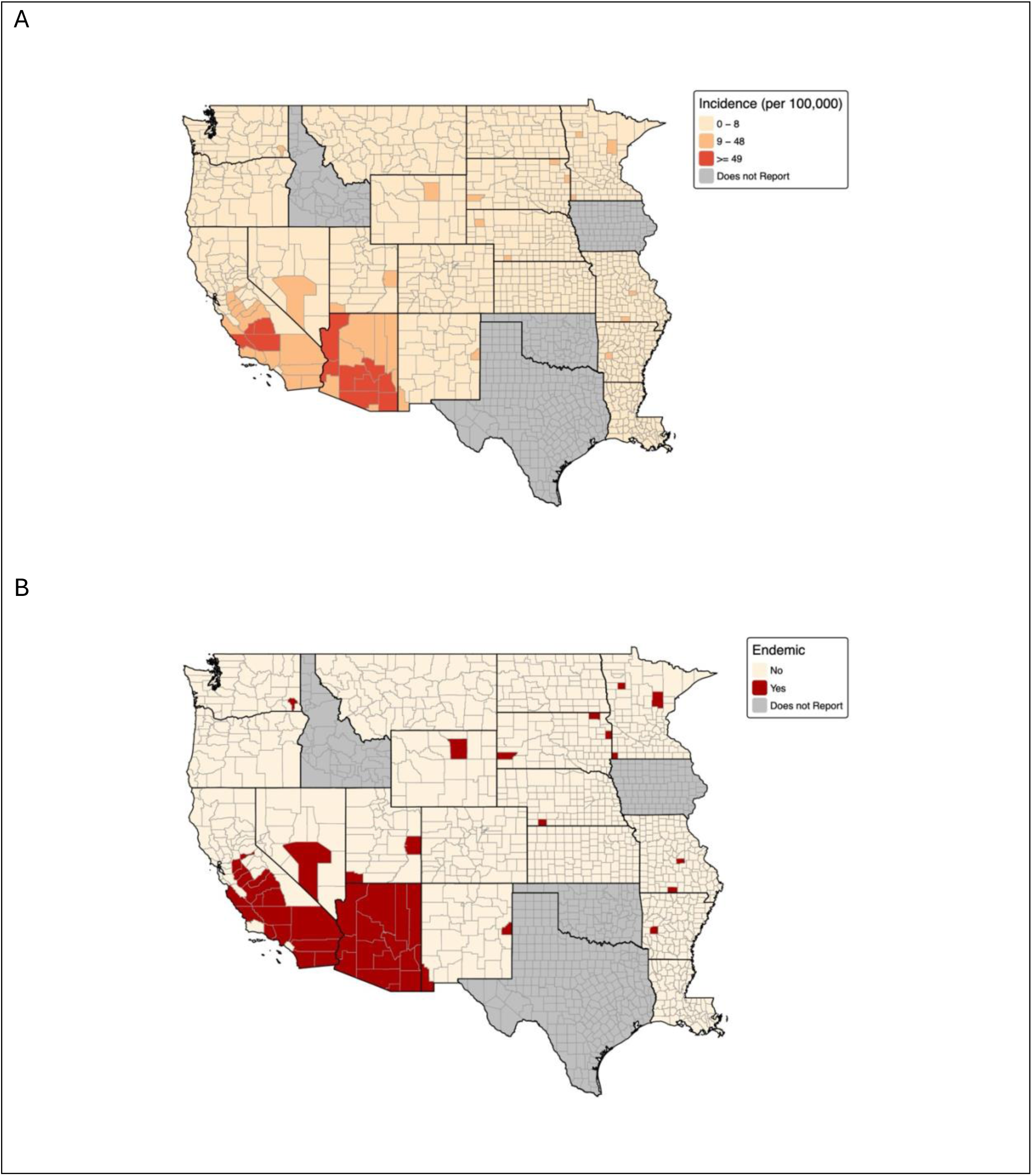
Coccidiomycosis incidence per 100,000 individuals at the county level in 2022 (A), and counties which met the endemicity definition of 10 or more cases per 100,000 in 2022 (B). Valley fever case data was provided by the CDC. Does not report means the county is in a state where Coccidiomycosis is not reported to the CDC.

Endemic counties were concentrated in the southwest, and incidence was highest in Kern County, California (265 cases per 100,000) and Pinal County, Arizona (193 cases per 100,000). Forty nine of 1,501 counties in our study area met CDC criteria for endemicity (incidence >=10 per 100,000) (Fig. 1b). While most endemic counties are in Arizona (15 counties) and the Central Valley of California (17), Utah, New Mexico, and Nevada also report endemic regions. We also observed counties meeting the CDC’s criteria for endemicity in Minnesota and Missouri, further east than the expected range of

*Coccidioides*.

To explore spatial variation in human case reports, we fit a spatial INLA model with no covariates which accounts for spatial autocorrelation in case reports and serves as a baseline for evaluating the effect of ecological covariates. The spatial-only model showed substantial county-level variation, indicating structured spatial heterogeneity in endemicity that environmental covariates might explain..

To assess whether the distribution *Coccidioides* reservoirs is associated with reported coccidioidomycosis, we first mapped the distributions of all previously described confirmed small mammal reservoirs of *Coccidioides* (Fig. 2). Counties with high incidence of reported disease also have the highest number of potential reservoir species (Fig. 2). Fifteen counties in Arizona, and 19 counties in California, have more than ten endemic small mammal reservoirs. *Coccidioides* reservoirs are also found in parts of Utah, Colorado, New Mexico, and Texas.

**Figure 2:**
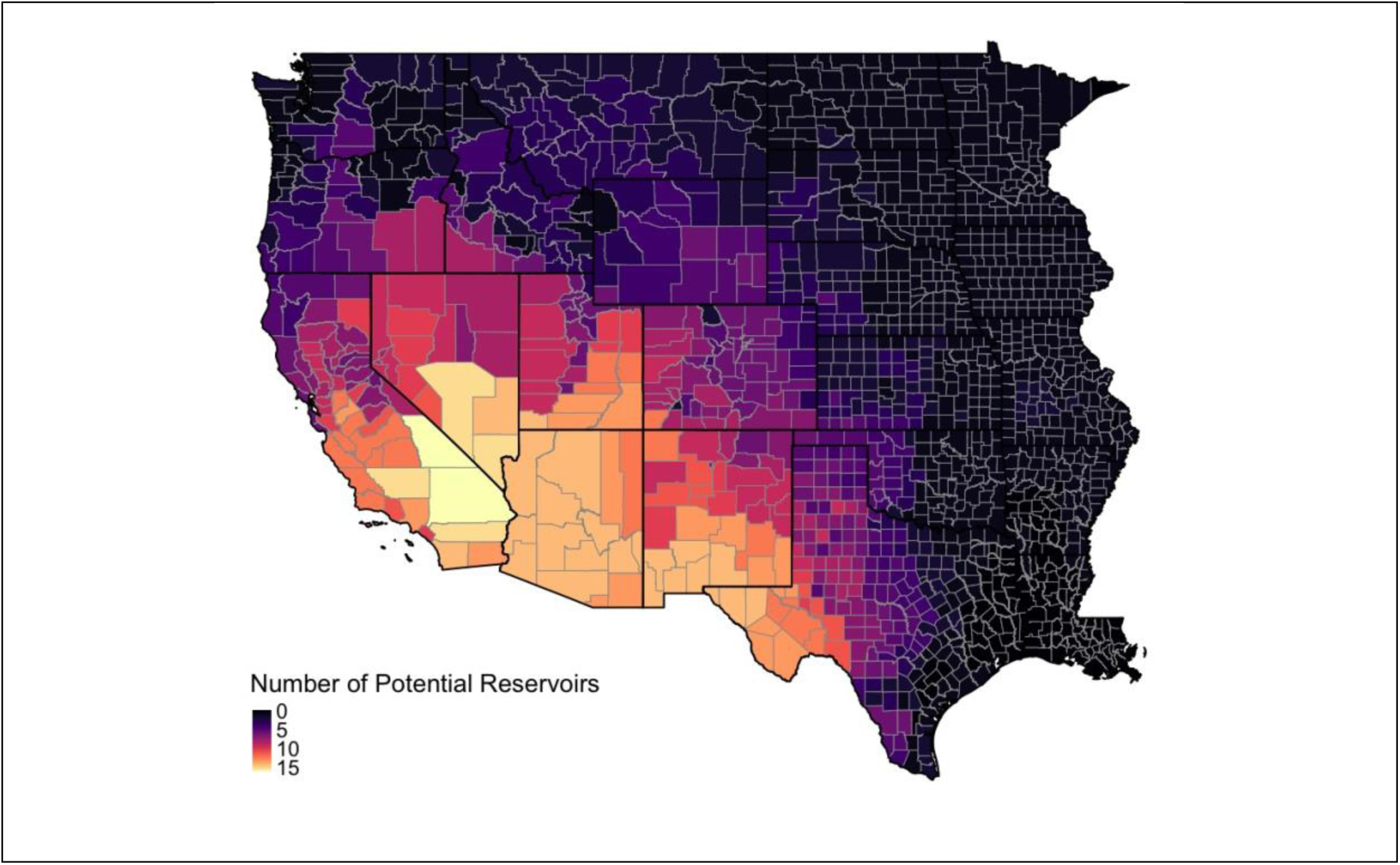
Number of potential *Coccidioides* reservoir mammal species predicted to be present in each western U.S. county, based on species distribution model overlap.

We then fit a spatial INLA model to predict county-level coccidioidomycosis endemicity including the number of locally endemic *Coccidioides* reservoir species, VPD, and mean soil moisture among others. Model predictions of coccidioidomycosis endemicity was largely consistent with reported cases, with endemic counties concentrated in Arizona and the Central Valley, California (Fig. 4a). Thirty-one counties had >50% probability of endemicity, clustered in Arizona and California’s Central Valley. The highest predicted probability was 96% in Maricopa County, Arizona.

Presence of *Coccidioides* reservoir species was the strongest predictor of county endemicity (log-odds ratio per SD = 1.702; 95% CI: 1.060-2.419) (Fig. 3a). Other environmental variables, including vegetation, soil moisture, soil bulk density, maximum temperature, and land cover category were not significantly associated with endemicity (Fig. 3a). We performed additional sensitivity analysis across endemicity thresholds to examine the robustness of the positive association between reservoir presence and endemicity. Reservoir presence remained significant when the endemicity threshold was reduced to 5 cases per 100,000 (LOR: 1.217; 95% CI: 0.703-1.746) and increased to 15 cases per 100,000 (LOR: 1.674; 95% CI: 0.792-2.461).

**Figure 3:**
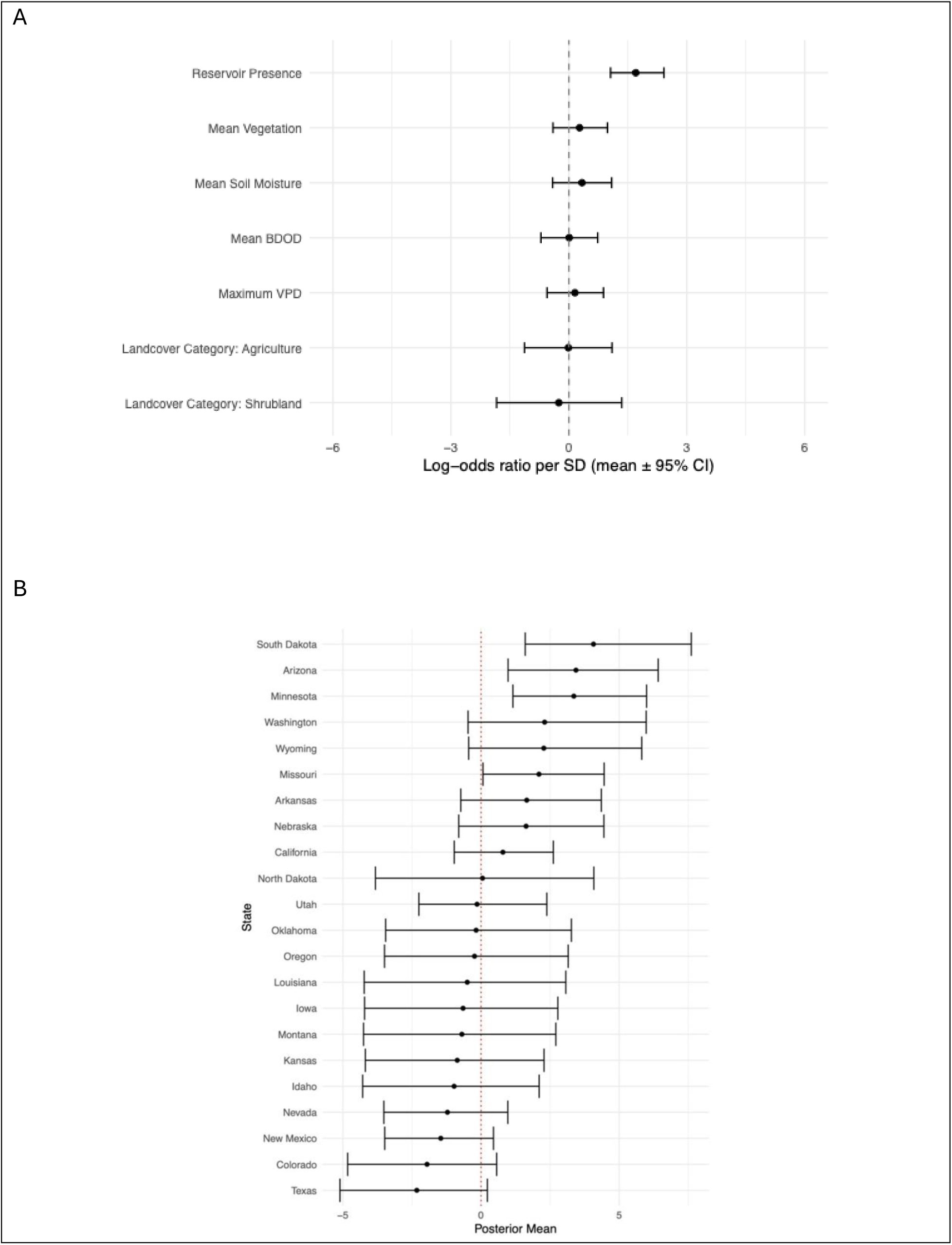
Log-odds ratio means for model fixed effects and 95% C.I. (A), and posterior mean of model state I.I.D. effect (B).

**Figure 4:**
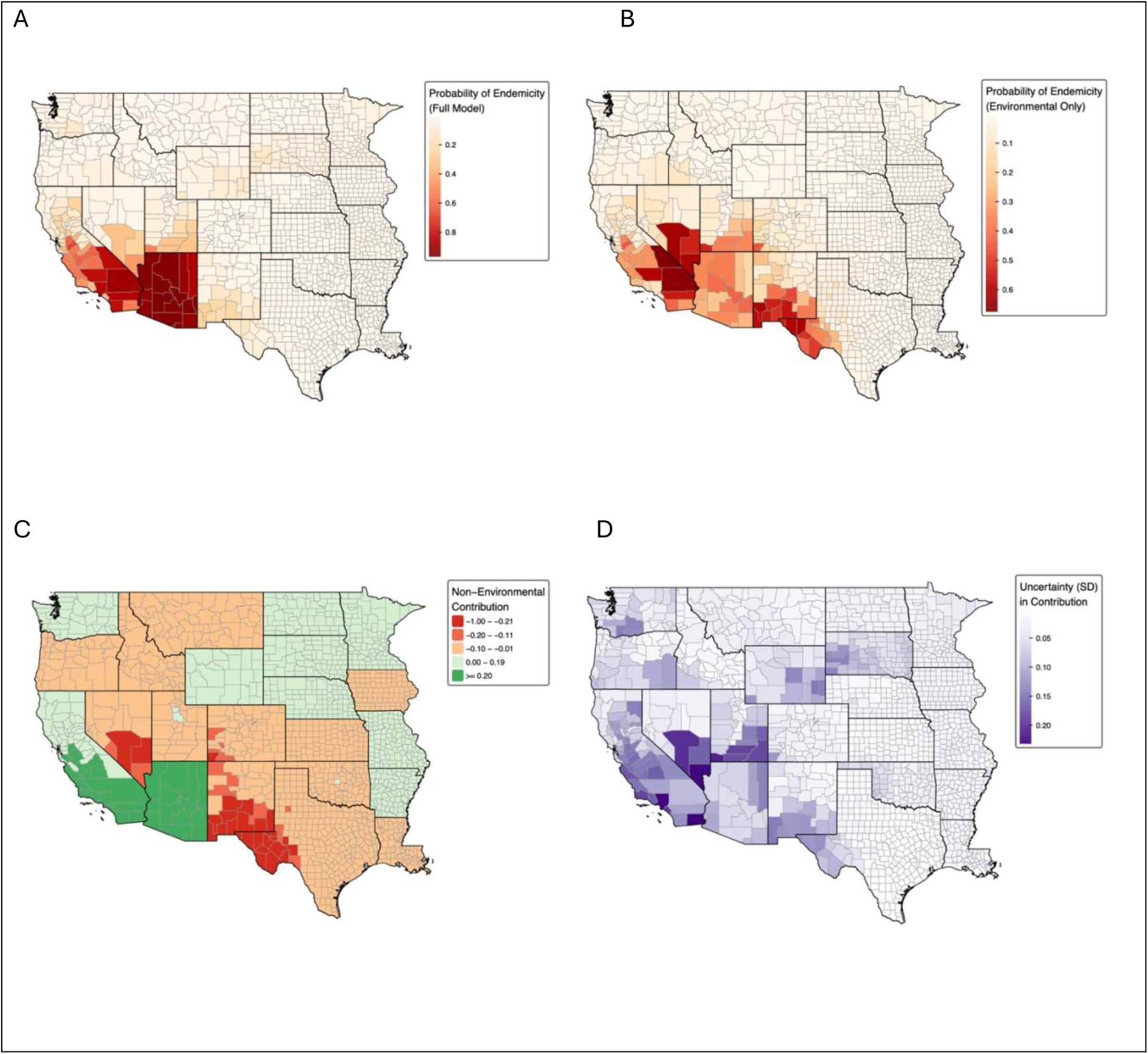
Modeled probability of endemicity of reported Coccidioidomycosis disease from the full model (A), modeled probability of environmental endemicity using only environmental covariates (B), the difference between the two levels (C), and the standard deviation of the predicted difference in the probability of endemicity (D).

We compared multiple covariate specifications in order to examine the additional value of including reservoir range covariates in the model using WAIC. The model including mammal reservoir distributions achieved the lowest WAIC among covariate models (WAIC = 280.82), outperforming the model without reservoir predictors (change in WAIC = 32.71). This provides strong evidence that reservoir species presence contributes predictive power beyond other included ecological covariates.

State-level random effects captured differences in reporting probability not explained by ecological covariates (Fig. 3b). The Arizona random effect was significantly higher than zero (3.435, 95% CI: 0.977-6.411), indicating that cases were more likely to be reported here than expected by a model where reporting was constant across the Western US. South Dakota (4.07, 95% CI: 1.60-7.61) and Minnesota (3.358, 95% CI: 1.152-5.993) also had mean state random effects above zero (Fig. 3b). States with lower random effect values included New Mexico (−1.457, 95% CI: −3.486-0.450), Colorado (−1.955, 95% CI: −4.828-0.565), and Texas (−2.325, 95% CI: −5.107-0.229), though these did not differ significantly from zero.

The state-level random effect exhibited greater variation (SD = 1.38) than the county-level spatial effect (SD = 0.05), indicating state-level heterogeneity is primarily responsible for residual variation after accounting for environmental covariates. This is likely attributable to differences in reporting across states. The BYM2 parameter (φ = 0.29; 95% CI: 0.01–0.90) suggests predominantly unstructured variation, though the wide credible interval indicates substantial uncertainty about the balance of spatial structure. In our model unexplained county-level factors affecting endemicity do not exhibit strong spatial patterns beyond what is explained by environmental covariates.

Our hierarchical model structure also enabled us to make model predictions based solely on environmental covariates, excluding population, county spatial effects, and state reporting effects, which we interpreted as predictions of environmental endemicity of *Coccidioides*, regardless of human case reporting (Fig. 4b). The probability of environmental endemicity is, like human cases, concentrated in the Southwestern U.S. However, this model predicts a higher probability of environmental endemicity in southern Nevada, southern and eastern Utah, and southern New Mexico, and western Texas.

We calculated the difference between predicted environmental endemicity and observed human case endemicity (Fig. 4c). Negative differences indicate counties where the fungus is predicted to be present but human cases are under-reported. Arizona and California’s Central Valley and Bay Area exhibited above-average reporting rates. Reporting rates are lower than the mean in southern Nevada, southern and eastern Utah, southern New Mexico, western Texas, and southwestern Colorado (Fig. 4c). There is substantial uncertainty in the disparity between predicted environmental and human case endemicity, as shown by the standard deviation of the difference in predicted probabilities (Fig. 4d). The greatest uncertainty occurred in northern Arizona and southern Utah.

## Discussion

We found that the presence of small mammal reservoirs of *Coccidioides* was the most important predictor of coccidioidomycosis endemicity. We also observed substantial differences in state-level reporting, suggesting that surveillance infrastructure influences the apparent distribution of disease.

Our findings have important public health implications. First, the observed discrepancies between environmental suitability and reported cases highlight gaps in surveillance and case recognition. States with low relative reporting rates or where coccidioidomycosis is not reportable may be underestimating their true disease burden, limiting their ability to allocate resources and raise awareness of coccidioidomycosis risk among the public and healthcare providers. Second, the strong relationship between small mammal reservoirs and endemicity supports the endozoan hypothesis, which suggests that burrowing mammals harbor *Coccidioides* and contribute to its persistence in soil^8^. This suggests that mammal distributions provide important information for both understanding the current range of *Coccidioides* and predicting changes in its distribution under shifting climate and land use patterns. One mechanism through which reservoir richness increases cases may be that in areas of high species richness, both specialist and generalist species are present. Generalists are more likely to be commensal with humans and spread disease. Generalist species opportunistically occupy burrows made by other species. In an area with low reservoir richness, multiple types of burrows are less likely to be occupied, potentially decreasing *Coccidioides* transmission^39^.

Our results build upon prior models of the distribution of coccidioidomycosis. Previous work has focused heavily on climatic and soil covariates. Prior modeling approaches have adopted occupancy models to account for imperfect reporting^21^. Our approach builds on an occupancy modeling framework by predicting risk effectively in unsampled areas. By integrating ecological information on potential reservoirs with human surveillance data, our models advance the capacity to disentangle environmental and reporting factors underlying of observed endemicity. Although environmental predictors such as soil moisture and temperature are important for fungal survival, their correlation with small mammal distributions complicates inference.

Our study has several possible limitations. Reported human case data are biased by under-diagnosis, under-reporting, and variation in testing practices across states, which may lead to misclassification of endemic counties. Information on mammalian reservoirs is also incomplete; for example, the apparent scarcity of reservoir species in Oregon and Washington could reflect biological reality or simply a lack of systematic screening of mammals in those regions. Incomplete sampling of small mammals across their range can lead to uncertain range maps Furthermore, many of the covariates included in our models are interdependent: small mammal ranges are shaped by environmental conditions, making it difficult to determine causality. Lastly, our study considers the distribution of *Coccidioides* in a single year. Temporal and seasonal changes in our covariates and Valley fever case reporting may add additional information to explain the role of mammal reservoirs in *Coccidioides* endemicity and may help generate more stable predictions of future Valley fever risk. Expanding our modeling approach to include a temporal component is an important future direction.

Enhanced surveillance systems in under-reporting states will be critical to improve recognition of Valley fever and to better define its true range. Future research should prioritize systematic surveys of small mammal populations across the western United States to clarify which species are competent reservoirs of *Coccidioides*. In addition, preserved samples in natural history collections and biorepositories could provide additional insight into the historical spread of *Coccidioides*^40,41^. Finally, longitudinal field studies and genomic approaches that link mammalian ecology, fungal persistence, and human infections are needed to establish causal pathways and refine predictive models.

Together, our findings underscore the importance of considering both ecological and surveillance in understanding the distribution of coccidioidomycosis. By incorporating small mammal reservoirs into models of endemicity, we provide stronger evidence for the endozoan hypothesis and highlight critical gaps in public health reporting. These results set the stage for more targeted ecological and epidemiological studies to inform prevention and control of Valley fever.

## Data Availability

All data produced in the present study are available upon reasonable request to the authors, and code to obtain environmental covariates and fit the INLA model are available at https://github.com/sussmanjacob/vf_distribution_modeling. County level Valley fever case counts are not publicly available.

## Acknowledgments

This work was supported by a Burroughs Wellcome Fund (Climate and Health Interdisciplinary Award to KSW, KD, and KP).

## Significance statement

Valley fever, or coccidioidomycosis, is an emerging fungal disease caused by the pathogen *Coccidioides*, with a geographic range that is not well understood. Maps of Valley fever risk primarily rely on human case data, overlooking potentially ecological influences like wildlife. This study demonstrates that the ranges of mammalian reservoirs of *Coccidioides* help predict the distribution of human infection risk. By integrating species distribution models with epidemiological data, this study also identifies reporting gaps in the Western United States where *Coccidioides* may be endemic but human cases are underreported. Our findings highlight areas where unknown Valley fever risk may be present and offers a framework for anticipating disease risk as environmental shifts occur.

