## Supplementary Information for "Wildlife hosts predict the distribution of reported coccidioidomycosis in the western United States"

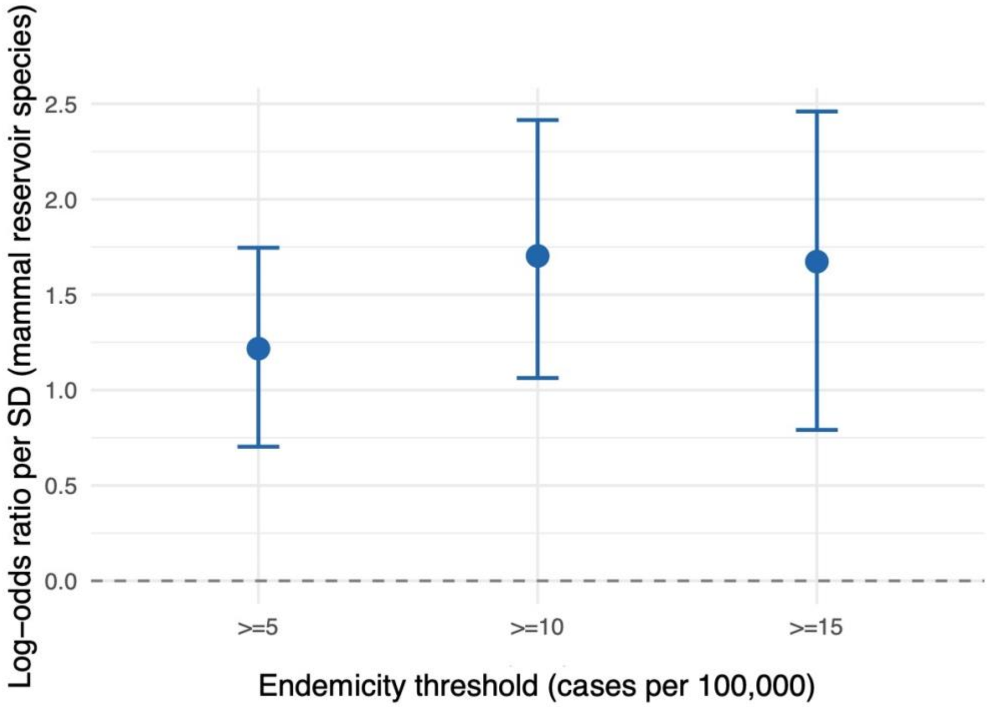


***Figure S1*****:** Mammal reservoir odds ratio from full model fitted to *Coccidioides* endemicity defined as >=5, >=10, and >= 15 cases per 100,000.

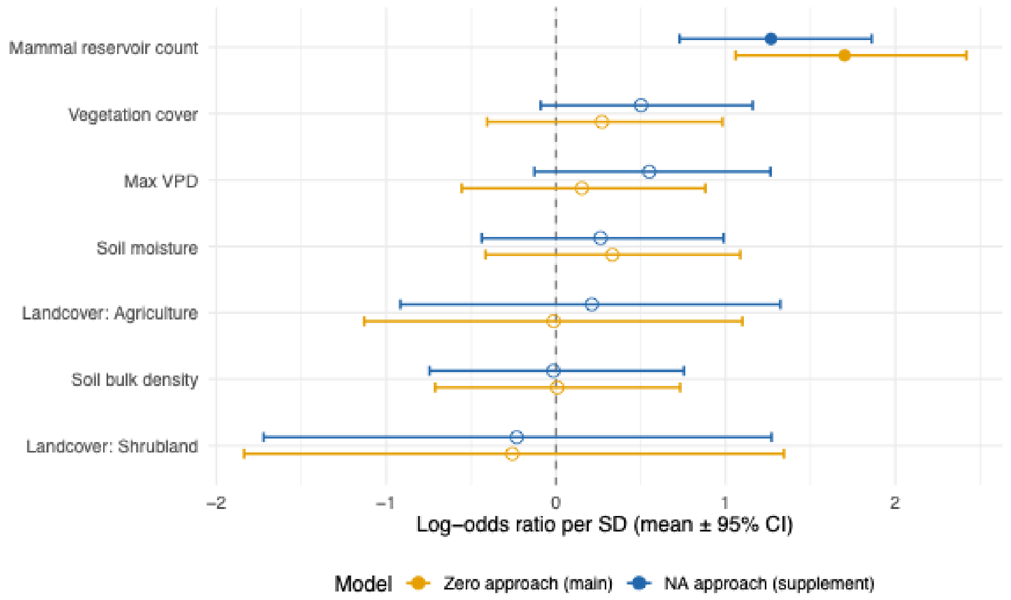


***Figure S2*****:** Log-odds ratio means for model fixed effects and 95% C.I, shown for analysis model, and supplemental model treating endemicity in states which do not report Valley fever as NA.

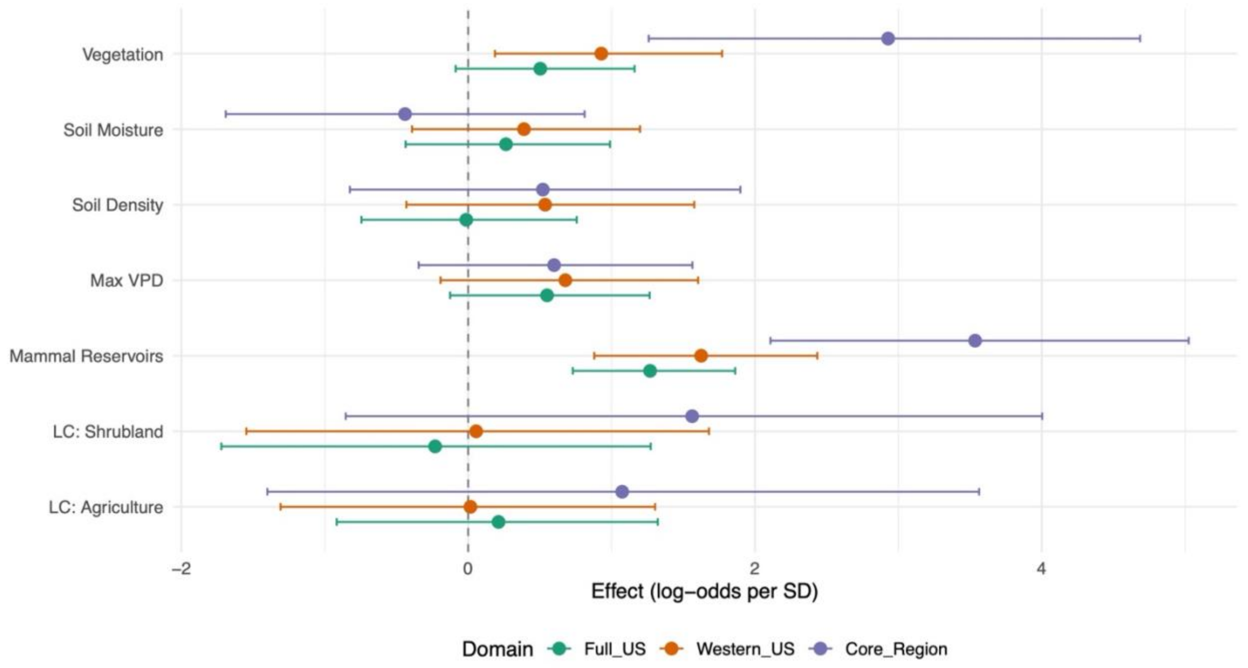


***Figure S3*****:** Covariate log-odds per standard deviation for analysis model, and additional models fit on different spatial domains (only states in the western U.S., and only states in the primary known *Coccidioides* endemic region)

A

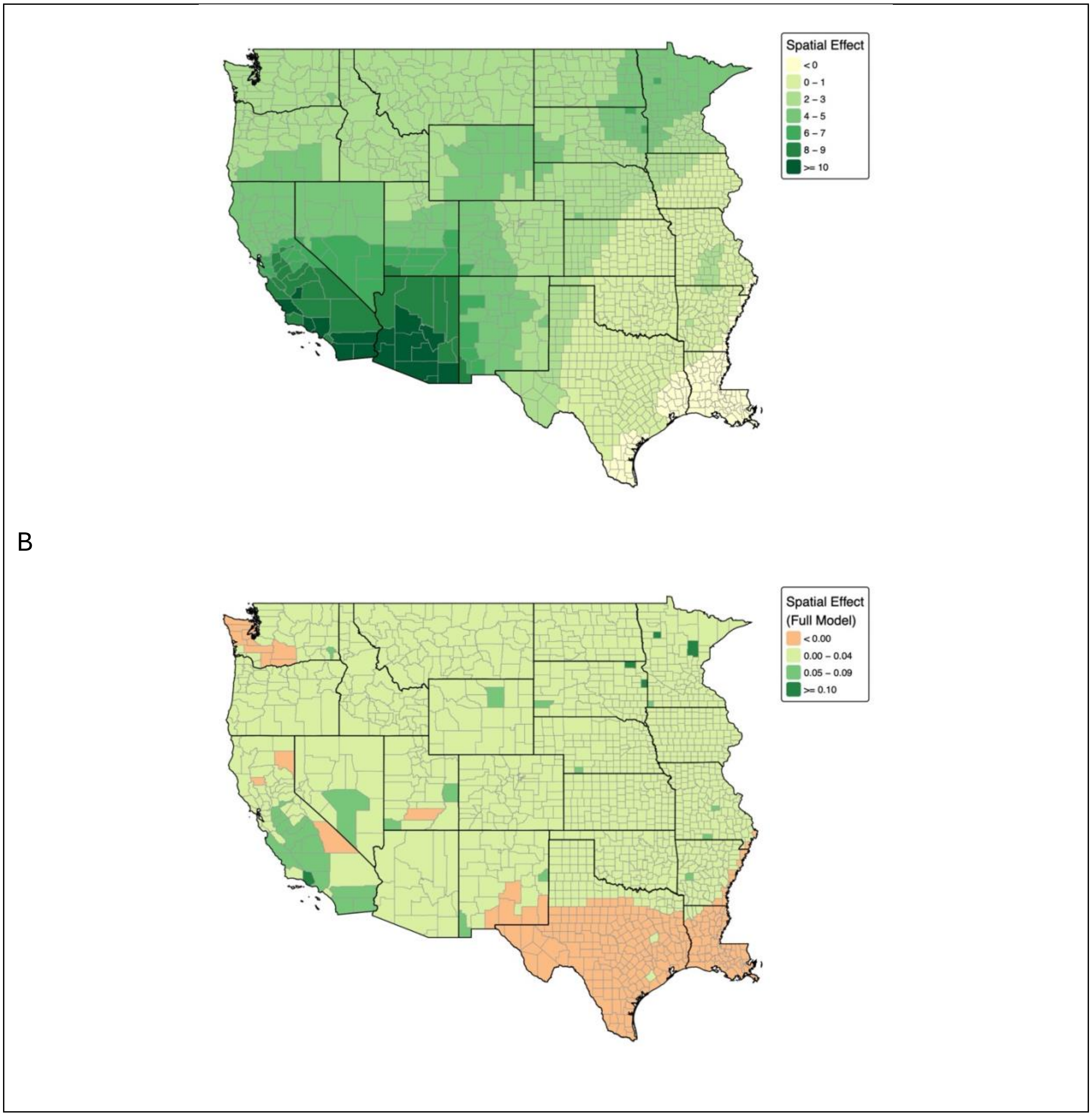


***Figure S4*****:** County level spatial effect for simple spatial model with no covariates (A), and full model (B).

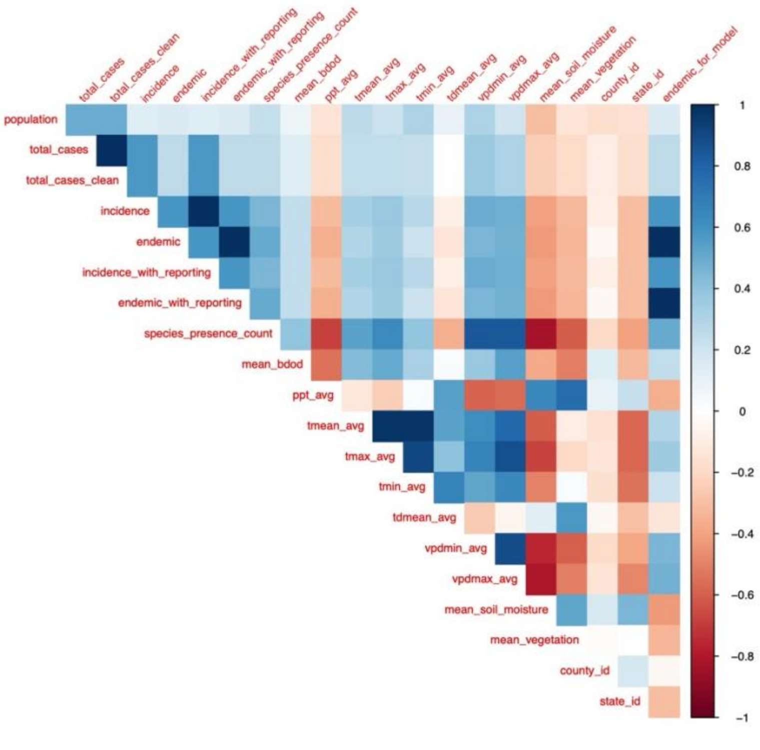


***Figure S5*:** Correlation heatmap for numeric covariates.

| Threshold (per 100k) | N endemic | % endemic | Log-OR | 95% CI | OR | OR 95% CI | Significant | WAIC |
|---|---|---|---|---|---|---|---|---|
| 5 | 87 | 2.8 | 1.217 | (0.703, 1.746) | 3.378 | (2.019, 5.734) | Yes | 458.21 |
| 10 | 53 | 1.7 | 1.703 | (1.063, 2.415) | 5.492 | (2.894, 11.194) | Yes | 289.23 |
| 15 | 34 | 1.1 | 1.673 | (0.791, 2.46) | 5.331 | (2.207, 11.709) | Yes | 233.11 |

***Table S1*: Model outcome and mammal reservoir log odds ratio for different endemic thresholds

| Category | States |
|---|---|
| Non-Reporting States | AK, CT, FL, GA, HI, ID, IL, IA, ME, MA, MS, NJ, NY, NC, OK, PA, SC, TN, TX, VT, VA, WV |
| Western US States | AZ, CA, CO, ID, MT, NV, NM, OR, UT, WA, WY, KS, MN, NE, ND, SD, OK, TX, AR, LA, MO, IA |
| Core Region States | AZ, CA, NV, NM, UT |

***Table S2*: Region definitions used for analysis

| Model | Covariates (n) | WAIC | ΔWAIC | DIC | pD |
|---|---|---|---|---|---|
| VPD replaces tmax * | 6 | 280.82 | 0 | 284.26 | 28.6 |
| No vegetation | 5 | 282.26 | 1.44 | 288.14 | 29 |
| No land cover | 5 | 283.41 | 2.59 | 290.02 | 30.9 |
| Temperature + VPD | 7 | 284.01 | 3.19 | 289.11 | 31.4 |
| Soil focused | 5 | 284.15 | 3.33 | 290.65 | 31.5 |
| Climate only | 5 | 286.04 | 5.23 | 299.07 | 37.2 |
| Minimal (4 covariates) | 4 | 288.36 | 7.55 | 293.39 | 31.8 |
| Base (tmax) | 6 | 288.85 | 8.03 | 295.38 | 33.6 |
| Add precipitation | 7 | 302.65 | 21.83 | 317.73 | 46.5 |
| Full climate suite | 8 | 305.94 | 25.13 | 325.32 | 51 |
| No mammal reservoirs | 5 | 313.52 | 32.71 | 325.01 | 38 |

***Table S3*:** Covariate model comparison. Models ranked by WAIC. * = selected final model specification. All models include BYM2 county spatial effect, state-level iid random effect, and population. ΔWAIC relative to lowest WAIC model.

| Host | State | County | Author | Date | Diagnostic |
|---|---|---|---|---|---|
| *Ammospermophilus harrisii* | AZ | Cochise | Salazar-Hamm, et. al. | 9/26/22 | ITS2 Sequencing |
| *Chaetodipus penicillatus* | AZ | Cochise | Salazar-Hamm, et. al. | 9/26/22 | ITS2 Sequencing |
| *Dipodomys merriami* | AZ | Cochise | Salazar-Hamm, et. al. | 9/26/22 | ITS2 Sequencing |
| *Sylvilagus audubonii* | AZ | Cochise | Salazar-Hamm, et. al. | 9/26/22 | ITS2 Sequencing |
| *Chaetodipus penicillatus* | AZ | Maricopa | Salazar-Hamm, et. al. | 9/26/22 | ITS2 Sequencing |
| *Dipodomys merriami* | AZ | Maricopa | Salazar-Hamm, et. al. | 9/26/22 | ITS2 Sequencing |
| *Dipodomys heermanni* | AZ | Maricopa | Salazar-Hamm, et. al. | 9/26/22 | ITS2 Sequencing |
| *Perognathus amplus* | AZ | Maricopa | Salazar-Hamm, et. al. | 9/26/22 | ITS2 Sequencing |
| *Dipodomys heermanni* | CA | Kern | Salazar-Hamm, et. al. | 9/26/22 | ITS2 Sequencing |
| *Dipodomys nitratoides* | CA | Kern | Salazar-Hamm, et. al. | 9/26/22 | ITS2 Sequencing |
| *Onychomys torridus* | CA | Kern | Salazar-Hamm, et. al. | 9/26/22 | ITS2 Sequencing |
| *Peromyscus maniculatus* | CA | Kern | Salazar-Hamm, et. al. | 9/26/22 | ITS2 Sequencing |
| *Thomomys bottae* | NM | Catron | Salazar-Hamm, et. al. | 9/26/22 | ITS2 Sequencing |
| *Chaetodipus intermedius* | NM | Sierra | Salazar-Hamm, et. al. | 9/26/22 | ITS2 Sequencing |
| *Dipodomys merriami* | NM | Sierra | Salazar-Hamm, et. al. | 9/26/22 | ITS2 Sequencing |
| *Neotoma albigula* | NM | Sierra | Salazar-Hamm, et. al. | 9/26/22 | ITS2 Sequencing |
| *Neotoma stephensi* | NM | Sierra | Salazar-Hamm, et. al. | 9/26/22 | ITS2 Sequencing |
| *Otospermophilus variegatus* | NM | Sierra | Salazar-Hamm, et. al. | 9/26/22 | ITS2 Sequencing |
| *Peromyscus boylii* | NM | Sierra | Salazar-Hamm, et. al. | 9/26/22 | ITS2 Sequencing |
| *Neotoma albigula* | NM | Socorro | Salazar-Hamm, et. al. | 9/26/22 | ITS2 Sequencing |
| *Peromyscus maniculatus* | | | Reyes-Montes, et. al. | 10/10/16 | Post-mortem (serology) |
| *Neotoma lepida* | | | Reyes-Montes, et. al. | 10/10/16 | Post-mortem (serology) |

| | | | | | |
|---|---|---|---|---|---|
| *Perognathus intermedius* | | | C. W. Emmons | 1/1/43 | Culture |
| *P. penicillatus* | | | C. W. Emmons | 1/1/43 | Culture |
| *P. baileyi* | | | C. W. Emmons | 1/1/43 | Culture |
| *Dipodomys merriami* | | | C. W. Emmons | 1/1/43 | Culture |

***Table S4*: Potential reservoir hosts included as mammal reservoirs. For each species the source and method of reported *Coccidioides* positivity is listed.
